# Long-read sequencing and benchmarking of saliva against blood for genomic variant detection in autism

**DOI:** 10.64898/2026.10.09.26365004

**Authors:** Yang Sui, Gabriele Campos, Youngjun Kwon, Isaac Wong, Nidhi Koundinya, Katherine M. Munson, Kendra Hoekzema, Marcelo Ayllon, Tianyun Wang, Emily Neuhaus, Evan E. Eichler

## Abstract

Autism is highly heritable, yet many cases remain genetically unsolved even after short-read sequencing (SRS). Long-read sequencing (LRS) increases sensitivity for pathogenic variant discovery, especially for complex structural variants (SVs) in repetitive regions missed by SRS. However, LRS largely relies on blood-derived DNA; whether saliva can support genome-wide LRS at population scale remains to be seen. We report the first systematic evaluation of saliva-derived DNA for genome-wide LRS in autism, benchmarking 105 new saliva samples against 68 new blood samples and integrating 189 previously published LRS blood samples to expand the autism cohort and improve ultra-rare pathogenic variant discovery. A matched blood–saliva quad provided direct comparisons of variant detection between sample sources. Saliva samples contained substantially more nonhuman DNA than blood (mean 10% versus 0.005% of reads), reducing mean effective genome coverage from an estimated 28X to 24X after removal of nonhuman reads, compared with 34X in blood samples. Despite this reduction, saliva and blood samples yielded comparable numbers of validated (22,591 versus 23,064) and rare (209 versus 213) SVs per sample, which was also observed in the quad when matching coverage. Saliva LRS identified four new pathogenic and likely pathogenic variants missed by SRS. We report variants affecting *MECP2, DNMT3A, CHD8, LMBRD2, POGZ*, and *PHF3*; four additional candidates with potential relevance to autism; and an increase of diagnostic yield of 9-18%. Saliva-derived DNA support blood-comparable discovery of pathogenic variants missed by SRS, including SVs, with increased sequencing coverage compensating for nonhuman DNA contamination and reduced assembly contiguity, thus showing saliva DNA collection as a feasible strategy for large-scale LRS studies of patients and population-scale sequencing of cohorts.

## INTRODUCTION

Despite the widespread adoption of short-read sequencing (SRS) in clinical and research pipelines, pathogenic variants have not been discovered for the majority of patients. Diagnostic yields for autism (<30%) are markedly low^1–7^, reflecting both true genetic heterogeneity and a persistent technical blind spot: structural variants (SVs), tandem repeat (TR) variations, and variants embedded in repetitive regions or other complex regions are systematically under-ascertained by short reads. Long-read sequencing (LRS) directly addresses this blind spot by generating phased, near-complete genome assemblies and improving the detection sensitivity of *de novo* mutations (DNMs) and SVs, particularly in previously inaccessible genomic regions^8,9^. As a result, LRS has repeatedly recovered pathogenic variation invisible to SRS across rare disease and neurodevelopmental disorder (NDD) cohorts^10–13^.

In autism, LRS of 267 individuals from 63 families increased detection of gene-disrupting SVs and TRs by 33% and 38%, respectively, over SRS studies, and rare damaging variants accounted for 7.4% of autism cases^14^. LRS has also resolved complex genomic rearrangements in five autism families^12^, and our pangenome-based study of 51 unsolved families involving 189 individuals recovered variants missed by SRS studies, with a 5.9% pathogenic discovery rate^11^. Studies of NDDs and rare diseases show similar gains. Targeted LRS resolved all eight complex rearrangements and identified pathogenic or likely pathogenic variants in six of ten individuals lacking a diagnosis^15^. Similarly, LRS application added diagnoses in 7-10% of patients in short-read-negative NDD cohorts^16–18^. LRS efforts in disease cohorts typically spanning tens to a few hundred individuals have consistently demonstrated added diagnostic value over SRS yet have remained modest in scale, limiting statistical power to detect rare, large-effect variants and resolve genome-wide burden across variant classes^11^.

Most LRS studies use high-molecular-weight (HMW) DNA from blood or lymphoblastoid cell lines (LCLs). Blood is difficult to collect at scale in pediatric cohorts and remotely recruited families, whereas noninvasive, self-collected saliva is more practical, especially among children with developmental disabilities. Surveys of parents consistently show a preference for saliva over venipuncture^19^, while geneticists have preferred DNA obtained from peripheral lymphocytes, especially for LRS work^8,10–18,20,21^. To date, LRS studies using saliva have been few and have all relied on nanopore adaptive sampling for targeted recovery of select regions. Gamaarachchi et al. used iterative adaptive sampling to target the sequence and assembly of two complex disease loci, *FSHD1* and *MUC1-ADTKD*, from saliva^22^. Gocuk et al. applied adaptive sampling to enrich the X chromosome in blood, saliva, and buccal DNA from 16 female carriers of X-linked inherited retinal disease. Saliva DNA was of sufficient integrity to support both haplotype phasing and measure X-inactivation skewing^23^. Both studies increased coverage by adaptive sampling; neither compared genome-wide variant discovery between matched blood and saliva.

Here, leveraging a saliva-derived dataset from Simons Foundation Powering Autism Research for Knowledge (SPARK), we conducted a systematic study to directly compare genome-wide variant discovery between saliva- and blood-derived samples from unsolved autism families using PacBio high-fidelity (HiFi) LRS. We extended the analysis framework established in our previous LRS study^11^ to the recently released SPARK PacBio long-read whole-genome sequencing (lrWGS) pilot cohort and newly generated blood-derived LRS data. Within this integrated dataset, we compared saliva- and blood-derived LRS data using matched samples from the same autism multiplex quad and systematically characterized SVs, TRs, and DNMs, with a focus on variants that may have been missed by prior SRS analyses. Although saliva-derived DNA contain a higher proportion of nonhuman reads, we observe comparable genome-wide variant discovery between saliva- and blood-derived LRS data after adjusting for sequence coverage. Together, these results support saliva as a scalable source for HiFi LRS production and future large-scale sequencing studies.

## RESULTS

### Long-read data integration of 104 unsolved autism families

We generated a combined LRS resource of 362 samples from 104 autism families, integrating 173 previously unpublished samples (169 individuals) from 53 families and 189 samples from 51 previously studied families^11^ (**Fig. 1a**). The newly generated dataset included 16 quads from the Simons Simplex Collection (SSC), classified as unsolved according to the criteria used in Sui et al.^11^, based on prior Illumina whole-genome sequencing (WGS) studies^1,2^. These were sequenced from blood- or LCL-derived DNA using the PacBio HiFi platform. The remaining 37 families were from the SPARK PacBio lrWGS Pilot cohort and had been classified as unsolved based on prior Illumina whole-exome sequencing (WES), with no previously identified returnable autism variants or self-reported genetic diagnoses. Patients were selected with ≥3 phenotypic features to increase the likelihood of a returnable genetic result. Saliva DNA was either previously extracted at PreventionGenetics using a Chemagic magnetic-bead protocol (n = 95) or newly collected and extracted using the PacBio HMW Nanobind protocol (n = 13), with three samples processed using both methods. Saliva HMW DNA was enriched for fragments greater than 10 kbp before shearing to approximately 15 kbp for SMRTbell library preparation, whereas blood DNA libraries used alternative shearing and gel-based size selection to target a library size of ~20 kbp before sequencing on PacBio Revio instruments (**Methods**). To enable benchmarking of saliva-versus blood-derived LRS data, we collected fresh blood from one of these 37 SPARK families through Simons Research Match and generated LRS data using the same platform as for the SSC quads, providing paired saliva- and blood-derived LRS data from the same four individuals. Thus, the combined cohort included 52 simplex quads, 46 simplex trios, 1 multiplex quad with paired blood–saliva sequencing, 3 duos, and 2 singletons (**Fig. 1a, Dataset S1**).

**Figure 1.**
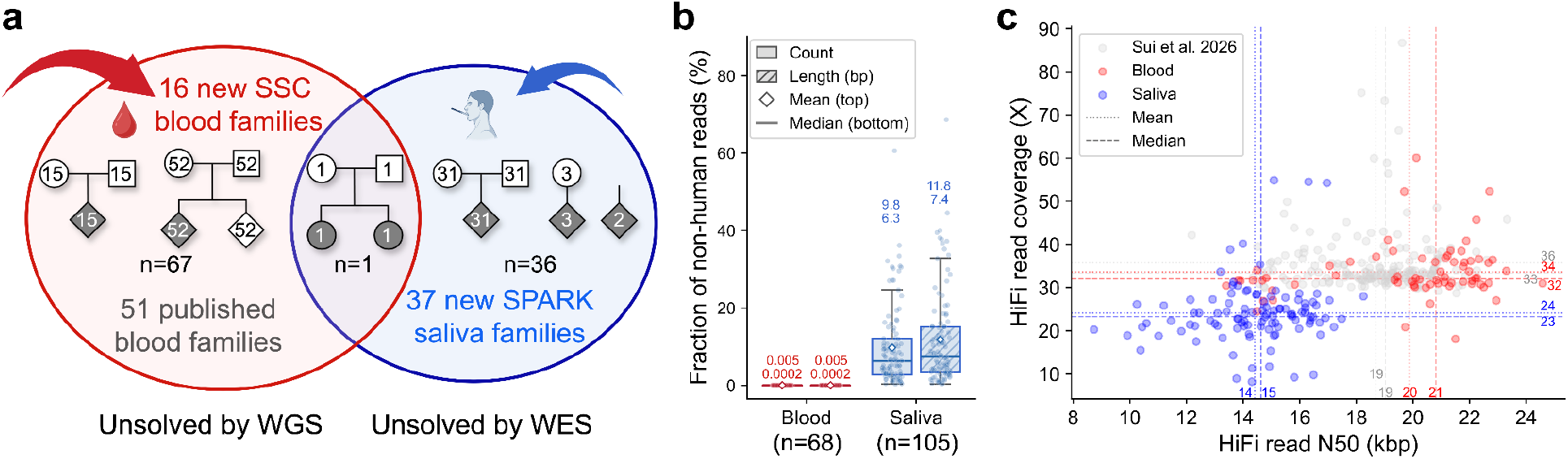
Overview of the combined long-read sequencing (LRS) resource of unsolved autism families. **a)** Schematic of the LRS study cohort and family pedigree structures. The integrated resource consists of 362 samples from 104 autism families, including previously unpublished LRS data from 53 families (169 individuals) compared to 51 families previously analyzed^11^. The new cohort includes 16 SSC quads classified as unsolved based on prior Illumina whole-genome sequencing (WGS) and 37 SPARK families classified as unsolved based on prior Illumina whole-exome sequencing (WES). One SPARK family was used as a control for benchmarking and DNA from blood and saliva from all four individuals was sequenced and analyzed. **b)** Box plots show the proportion of nonhuman reads identified by Kraken2^24^ (v2.1.3), measured by read counts and total read length (hatched), across 68 blood and 105 saliva samples. **c)** HiFi read N50 and genomic coverage per sample after removal of nonhuman reads. Dotted lines represent mean values, and dashed lines represent median values. Blood- and saliva-derived samples are shown in red and blue, respectively, and previously published data are shown in gray.

We assessed potential nonhuman contamination in all new LRS data using Kraken2^24^ (v2.1.3). Among the 68 newly generated blood samples, nonhuman reads were almost completely absent, with a mean fraction of 0.005% and a median of 0.0002% (0-0.1%) of reads, the corresponding fraction of total sequenced bases was virtually identical (**Fig. 1b**). In contrast, the 105 saliva samples show substantially greater contamination, with a mean fraction of 10% and a median of 6% of reads (0.3-61%), corresponding to a mean of 12% and a median of 7% of total sequenced bases (**Fig. 1b, Dataset S1**). After excluding these reads, the blood samples had a mean effective coverage of 34X (standard deviation, s.d. = 6), with a mean HiFi read N50 of 20 kbp (s.d. = 3), whereas the saliva samples had a mean effective coverage of 24X (s.d. = 8), reduced from an estimated 28X before removal of nonhuman reads, with a mean HiFi read N50 of 14 kbp (s.d. = 2; **Fig. 1c, Dataset S1**).

### Variant discovery in the discovery cohort

We applied an LRS variant discovery workflow established previously^9,11^ to the newly generated discovery cohort. Briefly, we generated genome assemblies from HiFi reads for each new sample using hifiasm^25^ (v0.25.0), producing haplotype-resolved (trio-binned) assemblies where parental WGS data (Illumina or PacBio HiFi) were available. Across the discovery cohort, assembly contiguity and consensus accuracy (PHRED quality value, QV) both increased with HiFi coverage (**Fig. 2a, b**). The 68 new blood samples achieved a mean contig N50 length of 47 Mbp (s.d. = 20), and a mean QV of 72 (s.d. = 1). In contrast, the 105 saliva-derived samples had substantially lower assembly contiguity, with a mean contig N50 of 17 Mbp (s.d. = 16), while maintaining a comparable mean consensus QV of 69 (s.d. = 2).

**Figure 2.**
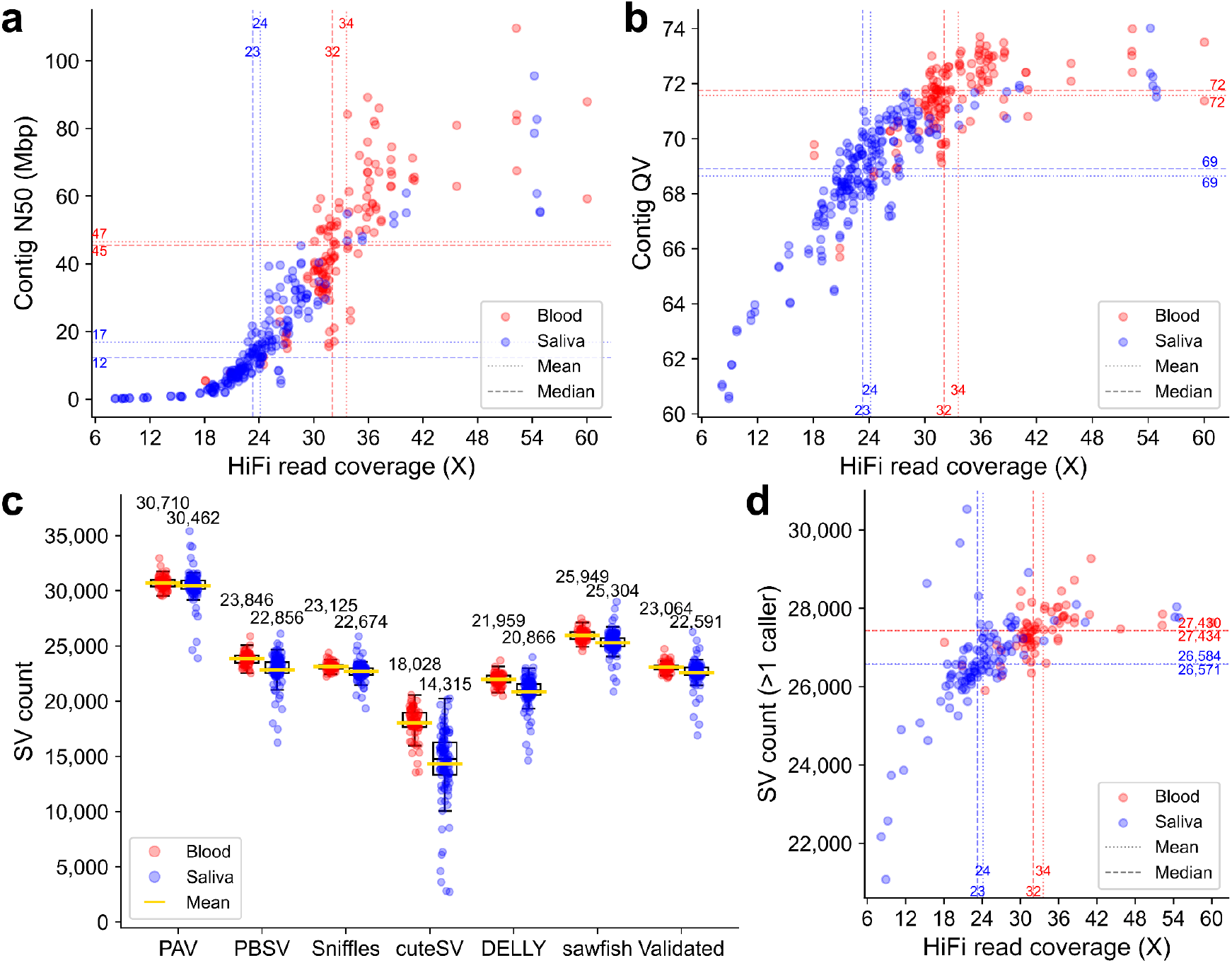
Long-read assembly and structural variant (SV) in the discovery cohort. **a-b)** Phased assembly N50 length (Mbp) (**a**) and sequence accuracy (QV, **b**) as a function of HiFi sequencing coverage in blood (red) and saliva (blue) samples. **c)** Number of SVs identified by each SV caller across the discovery cohort and the number of validated SVs per sample, defined as SVs identified by PAV and independently supported by at least one alignment-based caller. **d)** Relationship between effective HiFi sequencing coverage and the number of SVs supported by any two independent SV callers, irrespective of PAV support.

We discovered SVs as described previously^11^. We called SVs using phased assembly variant caller, PAV^26^ (v2.4.6), and five read-based alignment methods: PBSV (https://github.com/PacificBiosciences/pbsv, v2.9.0), Sniffles2^27^ (v2.3.2), cuteSV^28^ (v2.1.1), DELLY^29^ (v1.3.1), and sawfish^30^ (v0.12.4) (**Methods, Fig. 2c**). We considered PAV calls validated when independently supported by at least one alignment-based caller, as determined using Truvari^31^ (v5.2.0). Across the discovery cohort, an average of 22,777 validated SVs per sample were retained for downstream analysis. This yield was comparable to previously reported yields^11,32^ and differed only modestly between blood- and saliva-derived samples (23,064 versus 22,591 SVs per sample, respectively), with greater differences observed among low-coverage samples (**Dataset S1**).

When considering SVs supported by at least two independent callers irrespective of PAV support, blood samples consistently show a higher mean SV count than saliva samples (27,434 versus 26,584 per sample, respectively), reflecting the dependence of SV detection on sequencing coverage (**Fig. 2d**). We filtered the validated SV callset from 362 samples against 1,218 long-read population controls^32^ to identify rare SVs for downstream pathogenic candidate discovery (**Methods**, n = 617,488, **Dataset S2**). On average, we identify 164 rare SVs per child (n = 159, **Dataset S3**), corresponding to 0.72% of the validated SVs per child, compared with 202 rare SVs among 20,716 validated SVs (0.98%) in our previous study, which included 569 controls^11^. We then further excluded variants observed in 253 unaffected samples in the combined dataset or in gnomAD SVs^33^ (v4.1.0), with sex-aware filtering applied to the X and Y chromosomes. This filtering resulted in an average of 38 ultra-rare SVs per proband (n = 107, **Dataset S3**), representing 0.17% (38/22,777) of the validated SV calls per proband, which were subsequently annotated using comREG^11^ (v2.0.0) and evaluated for transmission.

In addition to SVs, we considered DNMs and used DeepVariant^34^ (v1.10.0) to generate callsets for all 362 samples, followed by joint genotyping using GLnexus^35^ (v1.4.1). We then overlapped the variants with the gnomAD^36^ v4.1.0 joint callset to filter out those variants observed in the general population (joint AC in exomes and genomes > 0) and excluded variants observed in 253 unaffected samples or located within homopolymer, gap, or centromeric regions. A total of 7,343 private variants specific to 102 probands were identified, with an average of 72 candidate DNMs per child after filtering with gnomAD, and annotated using the Ensembl Variant Effect Predictor (VEP^37^, v116.2) and comREG^11^ (v2.0.0) for functional prediction (**Dataset S4**).

### Benchmarking variant discovery between paired blood and saliva SPARK family

To assess the impact of tissue source on variant discovery, we randomly downsampled the saliva-derived HiFi reads from the benchmark family to match the blood-derived coverage for each individual (**Fig. 3a**). The family comprised two affected female children (designated p1 and p2) and both parents (saliva family identifier SF0036888; blood family identifier PM1001 in **Dataset S1**). HiFi reads are significantly shorter in saliva-derived samples compared to blood-derived DNA from the same donors, with mean read lengths of 12,086 ± 6,805 bp and 21,720 ± 5,698 bp, respectively, corresponding to a 44.4% reduction in mean read length in saliva (two-sided Welch’s t-test, p < 0.01, **Fig. 3a**). Downsampling consistently reduced assembly contiguity, but this reduction could be largely recovered with increased sequencing coverage (**Fig. 3b**).

**Figure 3.**
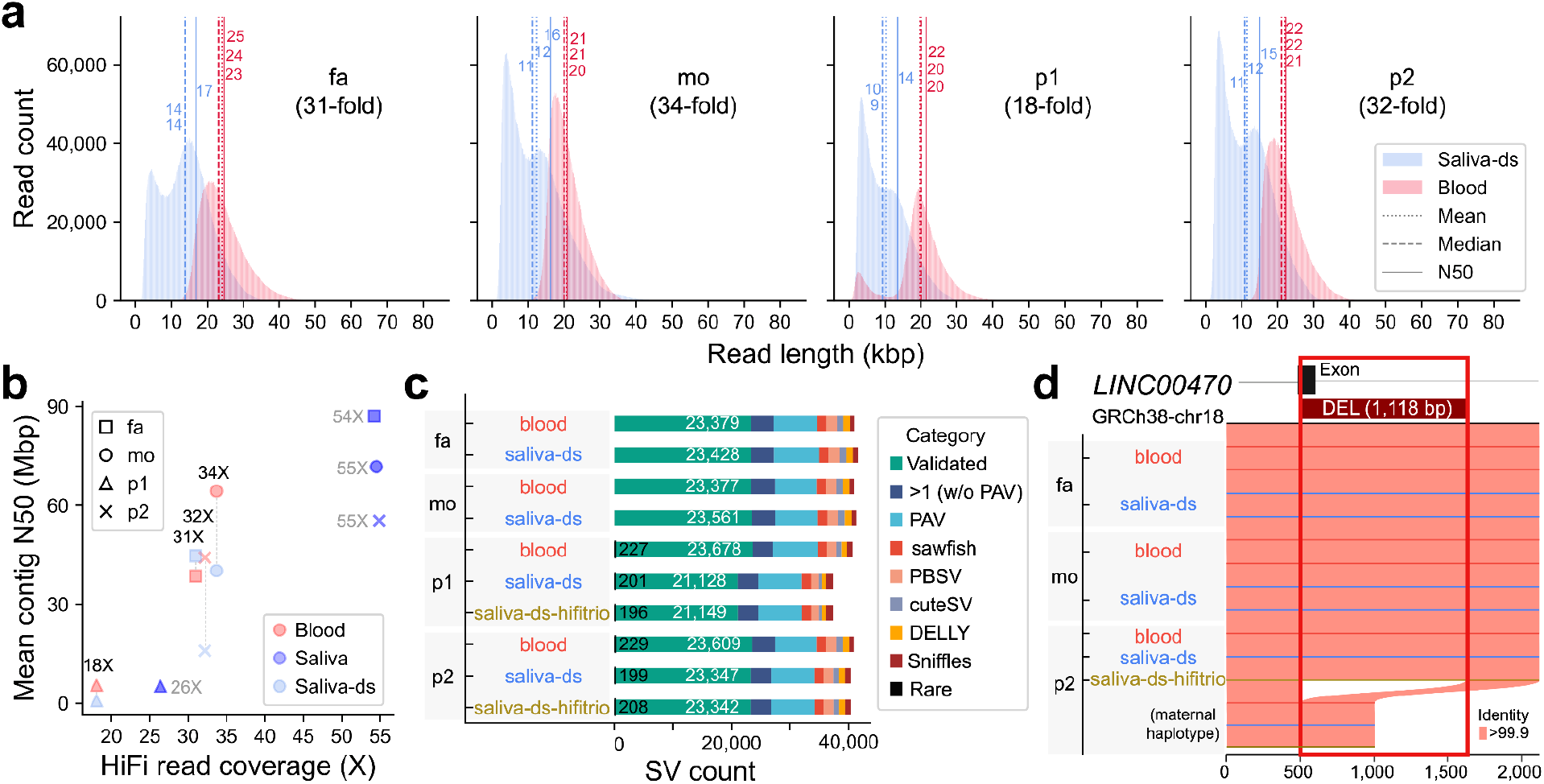
Benchmarking LRS performance and SV discovery using matched blood and saliva samples. **a)** HiFi read-length distributions for the four family members (father [fa], mother [mo], and two affected daughters [p1 and p2]) from saliva data randomly downsampled (Saliva-ds, light blue) to matched sequencing coverage with the corresponding blood samples (red). **b)** Contig N50 as a function of sequencing coverage for blood- (red) and saliva-derived assemblies before (blue) and after (light blue) saliva downsampling to matched coverage. **c)** Number of SV calls supported by different levels of caller concordance in blood and downsampled saliva samples, shown as stacked bars. Rare SVs not observed in the 1,218 controls are indicated in black within the validated SV set. HiFi-based parental phasing (saliva-ds-hifitrio, brown) was compared with Illumina-based parental phasing (saliva-ds, light blue) in the downsampled saliva data. **d)** A 1,118 bp *de novo* deletion identified as a candidate pathogenic variant in p2 in both blood and downsampled saliva data. The deletion removes part of the exon of *LINC00470* on the maternal haplotype. Each horizontal line represents a haplotype, and shading between haplotypes indicates sequence identity >99.9%.

We called DNMs and SVs independently from the downsampled saliva LRS data for all four family members as described above. Comparing blood and downsampled saliva LRS data while controlling for sequencing coverage (**Fig. 3c**), we detect no significant differences in overall SV counts (p > 0.125) or validated SVs (W = 3.0, p = 0.625, paired Wilcoxon signed-rank test). We additionally evaluated HiFi-based parental phasing for saliva samples lacking parental Illumina WGS data required for hifiasm phasing. Paired t-tests showed no significant difference between Illumina- and HiFi-based parental phasing (p > 0.126). Rare SV counts are likewise comparable among the three technical conditions, blood, downsampled saliva, and HiFi-phased saliva (227, 201, and 196 for p1; 229, 199, and 208 for p2, χ^2^ tests, p > 0.33, **Fig. 3c**).

Among the annotated rare SVs and DNMs, we identified a 1,118 bp *de novo* exonic deletion in a noncoding RNA gene, *LINC00470*, in a proband (p2) with neurological issues and language delay. The deletion was detected under all three technical conditions (**Fig. 3d, Table 1**) but missed by prior WES. As this is a novel candidate variant without sufficient evidence for pathogenic classification according to the American College of Medical Genetics and Genomics (ACMG) and the Association for Molecular Pathology (AMP) guidelines^38^, we considered it a variant of uncertain significance (VUS) and prioritized it for further investigation. Overall, these results demonstrate high concordance between blood- and saliva-derived sequencing data for profiling rare and potentially pathogenic variants.

**Table 1.** Pathogenic and candidate variants discovered by LRS.

| Sample | Sex | Gene | Chrom | Variant type | Variant class* | Source | SRS support |
| --- | --- | --- | --- | --- | --- | --- | --- |
| SF0483209_p1 | F | *MECP2* (SFARI1) | chrX | 35454 bp *de novo* DEL (exons) | Pathogenic | Saliva | No (WES) |
| SF0455838_p1 | F | *DNMT3A* (SFARI1) | chr2 | 302 bp *de novo* DEL (exons) | Pathogenic | Saliva | Yes (WES) |
| 13900_p1 | M | *CHD8* (SFARI1) | chr14 | 48 bp *de novo* splice-donor DEL | Pathogenic | Blood | No (WGS) |
| 14133_p1 | F | *PHF3* (SFARI1) | chr6 | 2 bp *de novo* frameshift DEL | Pathogenic | Blood | Yes (WGS) |
| SF0459673_p1 | F | *LMBRD2* (pLI0)<br>*ABHD17C* (pLI0.45) | chr5<br>chr15 | *de novo* missense (p.Arg483His)<br>*de novo* missense (p.Pro26Ala) | Pathogenic<br>VUS | Saliva | Yes (WES) |
| SF0491034_p1 | F | *POGZ* (SFARI1) | chr1 | *de novo* missense (p.Gly1256Cys) | Likely pathogenic | Saliva | Yes (WES) |
| SF0036888_p2,<br>PM1001_p2** | F | *LINC00470* | chr18 | 1118 bp *de novo* DEL (exon) | VUS | Saliva,<br>blood | No (WES) |
| SF0454336_p1 | M | *TWF2* (pLI0.97) | chr3 | 3290 bp *de novo* DEL (exon) | VUS | Saliva | No (WES) |
| SF0431733_p1 | F | *PCYOX1L* (pLI0) | chr5 | *de novo* missense (p.Phe204Ile) | VUS | Saliva | Yes (WES) |
| SF0452874_p1 | M | *TNPO1* (pLI1)<br>*EYA4* (pLI0.99) | chr5<br>chr6 | *de novo* missense (p.Val477Met)<br>*de novo* missense (p.Thr522Arg) | VUS<br>VUS | Saliva | Yes (WES) |

\*Pathogenic, likely pathogenic, and VUS (variant of uncertain significance) were classified according to the American College of Medical Genetics and Genomics (ACMG) guidelines<sup>38</sup>.
\*\*This is the same affected sibling (p2) from the multiplex benchmark family.

### Pathogenic and candidate variants identified in the LRS cohort

Following variant annotation and benchmarking of salivary LRS, we systematically evaluated ultra-rare SVs, TR variation, and DNMs across 107 probands to identify and nominate variants with potential relevance to autism. Of the three pathogenic variants and eight VUS reported in Sui et al.^11^, two previously categorized as homozygous pathogenic candidates, located in the regulatory regions of *ARHGEF10* (SFARI2) and *NCAPH2* (SFARI3), were excluded following filtering against the expanded population control set. In addition, we identified five novel pathogenic variants, one likely pathogenic variant, and four VUS, including the previously described 1,118 bp deletion in *LINC00470* identified from the multiplex benchmark family (**Table 1**). Among probands previously unsolved by short-read WGS, we identified pathogenic or candidate variants in 11 of 67 probands, including five pathogenic variants. Among those previously unsolved by short-read WES, we identified pathogenic or candidate variants in eight of 38 probands, including three pathogenic and one likely pathogenic variant. Thus, LRS provided an additional diagnostic yield of 7-19% for cases previously unsolved by WGS and 11-21% for those previously unsolved by WES, depending on whether candidate variants were included, corresponding to 9-18% when the two groups were combined (**Dataset S5**).

For example, we identify loss-of-function alleles, such as a pathogenic 35,454 bp *de novo* deletion that removes three exons of *MECP2* in a female proband with neurological issues, language delay, seizures/epilepsy, and motor delay (SF0483209_p1) (**Fig. 4a**). *MECP2*, the RETT syndrome gene, encodes a dosage-sensitive gene encoding methyl-CpG binding protein 2, a chromatin-associated protein that regulates gene expression and plays an essential role in neurodevelopment and neuronal function^39^. This large deletion was not identified by prior WES. We also identified a 302 bp *de novo* deletion in a SFARI score 1 gene, *DNMT3A*, in a female proband with attention-deficit/hyperactivity disorder (ADHD), macrocephaly, deafness/hearing loss, strabismus, polydactyly, learning disability, social anxiety disorder/social phobia, and cognitive impairment (SF0455838_p1), disrupting two exons of *DNMT3A* (**Fig. 4b**). *DNMT3A* encodes DNA methyltransferase 3A and has been established as a monoallelic autosomal gene. After discovery, we find that the deletion could be genotyped using Kanpig^40^ (v2.0.2) in the corresponding Illumina WES data; however, it was not originally identified in the prior short-read WES analysis.

**Figure 4.**
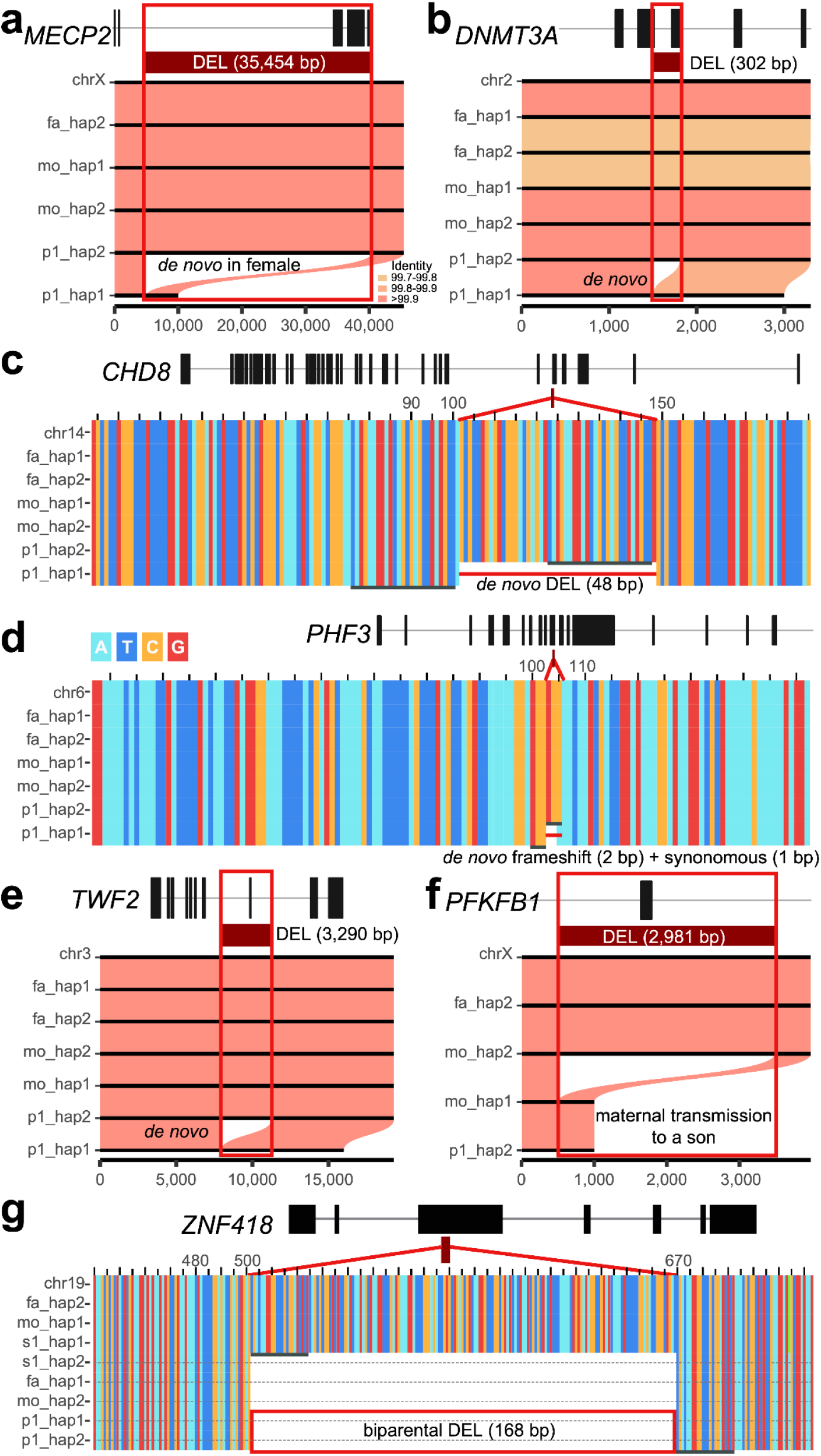
Pathogenic and candidate variants identified by LRS. **a)** A 35,454 bp *de novo* deletion in female SF0483209_p1 that deletes three exons of *MECP2*. Black lines represent haplotypes of samples in the autism family, with shading indicating sequence identity. **b)** A 302 bp *de novo* deletion in SF0455838_p1 that deletes exons of *DNMT3A*. **c)** A 48 bp *de novo* splice-donor deletion disrupting an exon of *CHD8* in 13900_p1. Multiple-sequence alignment (MSA) shows sequence-level evidence supporting the variant. Gray lines indicate repetitive sequences flanking the variant. **d)** A 2 bp *de novo* frameshift variant (p.Ala1081GlnfsTer2) and a nearby synonymous variant on the right (c.3243C>A, p.Ala1081%3D) affecting *PHF3* in 14133_p1. **e)** A 3,290 bp *de novo* deletion in SF0454336_p1 removing an exon of *TWF2*. **f)** A 2,981 bp deletion transmitted from the mother to her son (SF0441669_p1), deleting an exon of *PFKFB1*. **g)** A 168 bp biparental deletion in 11964_p1 inherited from both parents and deleting part of an exon of *ZNF418*.

A 48 bp *de novo* deletion (c.1593_1601+38del, autosomal dominant) affecting a splice-donor site of a SFARI score 1 gene, *CHD8*, was identified in a male proband with seizures, mixed expressive/receptive language disorder, speech delay, and nonverbal learning disorder (13900_p1, **Fig. 4c**). *CHD8* encodes chromodomain helicase DNA-binding protein 8 and has been established as a monoallelic autosomal gene with strong association with autism^41^. Of note, only three of 23 short reads (13%) supported the presence of the deletion, resulting in strong allelic imbalance that would likely cause the variant to be excluded by standard allele-balance filtering, whereas 18 of 32 HiFi reads (56%) supported the deletion. A 2 bp *de novo* frameshift deletion (c.3241_3242del, p.Ala1081GlnfsTer2, autosomal dominant) in *PHF3*, a SFARI score 1 gene encoding PHD-finger protein 3, a transcriptional regulator involved in neuronal gene regulation, was identified in a female proband with mixed expressive/receptive language disorder, pragmatics language disorder, and speech delay (14133_p1, **Fig. 4d**) and is predicted to produce a severely truncated protein of 1,082 amino acids, compared with the full-length 2,039-amino-acid protein and was classified as pathogenic according to ACMG guidelines. This DNM was missed in earlier analyses with SRS data^1,2^; however, examination of the previous SRS callsets confirms the presence of the variant, which was represented as two adjacent calls (chr6:63703543:CCG>C, frameshift and chr6:63703547:C>A, synonymous), corresponding to the same haplotype-level change (chr6:63703545:GCC>A, **Fig. 4d**), likely resulting in the complex allele being missed during subsequent variant filtering and annotation. Notably, the same filtering issues affected the previous LRS analysis of both the *CHD8* (**Fig. 4c**) and *PHF3* (**Fig. 4d**) variants, resulting in their omission from the candidate variant set; instead, a *de novo* promoter variant in *DDX3X* was prioritized as a candidate in 14133_p1^9,11^.

We also discovered a *de novo* pathogenic missense variant in *LMBRD2* (c.1448G>A, p.Arg483His, autosomal dominant) was identified in a female proband (SF0459673_p1) and was classified as pathogenic according to ACMG guidelines. *LMBRD2* (pLI = 0) encodes LMBR1 domain containing 2 protein, which is a widely expressed transmembrane protein implicated in adrenergic receptor signaling. The same missense variant has previously been reported as a recurrent DNM in three individuals with developmental delay (DD), intellectual disability (ID), and other neurodevelopmental features^42^. This proband presents with pervasive developmental disorder-not otherwise specified (PDD-NOS), ADHD, depression/dysthymia, obsessive-compulsive disorder (OCD), language delay, ID, cognitive impairment, global DD (IQ 40-54), social communication disorder, speech articulation problems, and neurological issues but also carried another *de novo* missense variant (c.76C>G, p.Pro26Ala) in *ABHD17C* (pLI = 0.45), which encodes an alpha/beta hydrolase that participates in protein depalmitoylation and regulates neuronal signaling networks involved in synaptic function and brain development^43^. We detected both variants in prior WES data and are uncertain why they were not called. (**Dataset S5**). In addition, we also identified a *de novo* likely pathogenic missense variant (c.3766G>T, p.Gly1256Cys, autosomal dominant) in a SFARI score 1 gene, *POGZ*, in a female proband with anxiety disorder, bipolar disorder, motor delay, language delay, learning disability, and speech articulation problems (SF0491034_p1). *POGZ* encodes the Pogo transposable element with zinc finger domain protein, a chromatin regulator and transcription factor involved in gene regulation and brain development^44^.

Among the variants of potential relevance to autism (i.e., VUS), we identified a 3,290 bp *de novo* deletion in *TWF2* (pLI = 0.97) in a male proband with ADHD, anxiety disorder, macrocephaly, cognitive impairment, motor delay, learning disability, and social communication disorder (SF0454336_p1, **Fig. 4e**) that deleted an exon of *TWF2*, which encodes Twinfilin-2 and has been implicated in modulation of synaptic structure^45^ and positive regulation of neuron projection development^46^. The deletion could not be genotyped by Kanpig using the corresponding WES data. A *de novo* missense variant (c.610T>A, p.Phe204Ile) in *PCYOX1L* (pLI = 0) was identified in proband SF0431733_p1, a female with ADHD, language delay, seizures/epilepsy, motor delay, ID, cognitive impairment, global DD, learning disability, and neurological issues. The variant was predicted to be deleterious by SIFT, PolyPhen-2, and AlphaMissense. *PCYOX1L* encodes prenylcysteine oxidase 1-like protein, a secreted protein implicated in regulating plasma-membrane surface expression of the synaptic ion channel ASIC1a, and genetic deletion of *PCYOX1L* has been reported to impair hippocampal long-term potentiation^47^. *PCYOX1L* is not currently listed in the SFARI Gene database but has recently been nominated as an emerging putative autism risk gene by Han et al.^48^.

In a male proband (SF0452874_p1), we identified two independent *de novo* missense variants, both computationally predicted to be damaging by SIFT, PolyPhen-2, and AlphaMissense, in two loss-of-function-intolerant genes: *TNPO1* (pLI = 1; c.1429G>A, p.Val477Met) and *EYA4* (pLI = 0.99; c.1565C>G, p.Thr522Arg). *TNPO1* (Transportin-1) is highly expressed in neurons and mediates nuclear import of numerous RNA-binding proteins^49,50^ and is required for normal neuronal development and function^51^. Its paralog *TNPO2* causes a defined autosomal dominant NDD marked by global DD, dysmorphic features, and neurologic and ophthalmologic abnormalities^52^. *EYA4* encodes a transcriptional coactivator/protein phosphatase acting with SIX and PAX family proteins during organ development^53^ and is an established autosomal dominant cause of progressive postlingual sensorineural hearing loss, occasionally accompanied by dilated cardiomyopathy^54,55^. The proband has cognitive impairment, language delay, ID, cognitive impairment, global DD, and learning disability, with no hearing loss or cardiac findings.

Finally, we also report three additional rare inherited and *de novo* variants in genes without established connections to autism, each noted for a specific inheritance pattern or functional prediction. A 2,981 bp deletion removes a single exon of *PFKFB1* in a male proband with short stature, motor delay, ID, cognitive impairment, global DD, learning disability, language delay, social communication disorder, and speech articulation problems (SF0441669_p1, **Fig. 4f**), which was inherited from his mother. Because *PFKFB1* is located on the X chromosome, the maternally inherited deletion leaves the proband hemizygous for the deleted exon. DECIPHER lists a separate hemizygous, maternally inherited *PFKFB1* variant (c.G>A, stop-gained) in an unrelated individual with abnormalities involving the head and neck, limbs, integument, musculoskeletal system, and nervous system; however, this variant is classified as a VUS. A *de novo* missense variant (c.350A>G, p.Tyr117Cys) in *ACTR6* (pLI = 0) was identified in a female proband with speech delay (13561_p1) and was predicted to be damaging by SIFT, PolyPhen-2, and AlphaMissense. A 168 bp deletion removing an exon of *ZNF418* in a female proband with speech delay (11964_p1, **Fig. 4g**) was biparentally inherited, consistent with a potentially homozygous deletion. None of these three genes are currently included in NDD gene databases, and additional genetic, functional, and clinical evidence will be needed to determine whether these variants are relevant to the observed phenotypes.

## DISCUSSION

Autism is highly heritable^56–58^, yet a substantial proportion of cases remains genetically unsolved based on previous large-scale short-read WES and WGS studies. Scaling family-based LRS discovery to the sample sizes needed to more comprehensively assess the missing heritability of autism requires leveraging larger existing genetic resources, including DNA obtained from saliva collections. In this study, we assessed the feasibility of LRS for variant discovery by integrating 362 long-read samples (257 blood and 105 saliva) from 104 unsolved autism families and benchmarked saliva against blood. Although saliva contained substantially more microbial DNA than blood (mean 10% versus 0.005% of reads), saliva-derived LRS recovered similar numbers of SVs, with approximately 22,591 validated SVs per sample. This yielded 209 ultra-rare SVs and 57 DNM candidates per proband after population- and family-based filtering for downstream pathogenic variant discovery. Similar results were observed in the matched blood–saliva family pair when adjusting for sequence coverage differences. Across the cohort, LRS increased the diagnostic yield by 9-18% in families unsolved by SRS, with comparable diagnostic yields for blood- and saliva-derived samples. This range is in line with the 7-17% estimated across rare-disease studies^17,18,20^ and the 7-33% reported in SRS-negative NDD cohorts^10,11,16,21^. Together, these findings support saliva as a viable source for genome-wide LRS studies.

Because there is, as of yet, no gnomAD database equivalent for long-read discovered genetic variants, an important consideration for discovery of ultra-rare or DNMs in these samples was to filter variants against population controls analyzed using the same callers and same sequence technologies^32^. To aid in this effort, we leverage in addition to these LRS population controls, 253 unaffected samples, and available SRS-based gnomAD data to exclude common variants in regions accessible by SRS. This procedure allowed us to prioritize approximately 110 variants (38 SVs and 72 DNMs) for pathogenic variant discovery among the affected children. This strategy discovered six novel pathogenic and likely pathogenic variants, as well as four additional candidate variants with potential relevance to autism for further functional testing. Among these candidates, a 1,118 bp *de novo* exonic deletion in *LINC00470* was reliably detected from DNA prepared from either blood or DNA. *LINC00470* has been implicated in PTEN regulation and neural development, while the deletion may also affect ELFN2-mediated glutamatergic signaling^59–62^, providing a biological rationale for further investigation. LRS of saliva DNA also identified a *de novo* exon deletion in *TWF2* and *de novo* missense variants in *PCYOX1L* and *TNPO1*, genes with emerging evidence for roles in synaptic, neuronal, or developmental function^45–47,51^. Notably, four variants were completely missed by SRS (**Table 1**) even after reanalysis owing to absent or poor SRS support, highlighting variants that would have been almost impossible to identify without LRS. Conversely, examination of the *CHD8* and *PHF3* variants identified by LRS revealed that complex local sequence contexts and differences in variant representation across calling algorithms can complicate variant calling and downstream filtering, underscoring the importance of considering sequence context when validating and interpreting candidate variants. While LRS may ultimately replace SRS as a one-stop genetic test^15,63–66^, in our study there was considerable value integrating LRS with systematic SRS reanalysis to validate events, especially more complex regions of the genome.

While saliva DNA show significantly increased contamination and overall shorter reads and contigs (**Figs. 1b-c, 2a, 3a**), our benchmark analysis of the blood–saliva matched quad revealed that these deficits can be compensated by increasing sequence coverage (**Fig. 3b**). Higher coverages not only improve contiguity of assembly but in some cases, surprisingly, exceed that of blood-derived DNA (**Fig. 3b**). Despite shorter read lengths, the high accuracy of HiFi drives assembly resulting in effectively large, phased contigs facilitating discovery of more complex repeat associated variants^25,26,67^ (**Fig. 2b**). Increasing saliva sequencing coverage, of course, comes with increased cost and we estimate a ~10% increase in SMRT cell usage to achieve the same coverage as blood DNA. With recent developments in SMRT cell reuse capability via PacBio SPRQ-Nx chemistry, such costs can be readily offset and we anticipate further reductions making HiFi WGS potentially the primary diagnostic tool for screening autism patients. Indeed, a recent clinical evaluation of ~1000 patients utilizing PacBio HiFi sequencing has shown it is ready to replace standard multi-step testing with a single-tier diagnostic test for rare genetic disease, and our analysis argues that saliva DNA may be a cheaper and more readily accessible substrate for this activity^68^.

An alternative LRS-based HiFi approach is ONT sequencing, which has long had flow cell reuse capability, though we did not evaluate this in this study. Specifically, ONT adaptive sampling provides an alternative strategy by rejecting non-target molecules during sequencing and thereby redirecting sequencing capacity toward human DNA, as demonstrated for saliva and other tissues in targeted sequencing studies^22,23^. However, ONT generally has lower per-base accuracy than PacBio HiFi, with errors particularly affecting homopolymers and short TRs. In addition, its utility for LRS of whole genomes obtained from saliva DNA remains largely unexplored. Saliva DNA may be less compatible with the ultra-long-read and Hi-C approaches used in some ONT-based assemblies for comprehensive phasing^22^. Nevertheless, PacBio HiFi and ONT offer different trade-offs with respect to contamination management, sequencing efficiency, read accuracy, and assembly and phasing capabilities. The optimal sequencing platform can therefore be selected based on the specific objectives of the research study.

The ability for saliva DNA to generate high-quality LRS data and assemblies resulting in comparable discovery of variants from blood-derived DNA is especially important for the autism research community. Surveys have shown that 92% of parents prefer saliva over blood collection due to the additional trauma associated with needle pricks and the need to on occasion restrain children^19^. SPARK consequently has shifted away from venipuncture to saliva collection to obtain the more than 50,000 families for genetic analyses^69^. These large-scale saliva collections therefore represent an important existing resource for extending LRS to the scale needed for autism genetics. Practically, if properly stored and quality controlled, such collections can be used for generating both SRS and LRS. The ability to leverage existing saliva collections for LRS could reduce barriers to recruitment and sequencing, particularly for autistic children and families for whom blood collection is challenging. This may also facilitate the inclusion of more complete family structures, which are particularly valuable for autism genetics because family-based analyses enable the identification and interpretation of both *de novo* and rare inherited variants. Thus, the compatibility of saliva DNA with LRS not only provides a practical alternative to blood collection but also creates an opportunity to extend LRS to existing and future large-scale autism cohorts, ultimately enabling more comprehensive characterization of the genetic architecture of autism.

## METHODS

### LRS data generation and QC

We generated PacBio HiFi sequencing data at the University of Washington for 68 blood-derived samples, including 58 samples with previously banked whole-blood DNA, 4 freshly collected whole-blood samples from the benchmark quad, and 6 SSC samples with low-input banked DNA that were supplemented with LCL-derived DNA. DNA from the four freshly collected whole-blood samples and the six LCL samples was extracted with the Monarch HMW DNA extraction kit for cells and blood (NEB T3050L) using an input of 500 uL whole blood or 5 million lymphoblastoid cells. We followed the manufacturer’s protocol, using 1000 rpm shaking speed during lysis. Following DNA extraction and QC, sequencing libraries were generated with the PacBio SMRTbell prep kit 3.0 and SMRTbell Adapter Index Plate 96A (102-182-700/102-009-200) per manufacturer’s protocol with the following modifications: DNA was sheared using the Megaruptor 3 (Hologic Diagenode P/N B06010003/E07010003) twice, at settings 28/30 or once, at setting 29, depending on input DNA intactness. After end repair, DNA damage repair, PacBio adapter ligation, and nuclease treatment, libraries were gel size-selected to remove short fragments and residual sequencing inhibitors on the Pippin HT (Sage Science P/N HTP0001/HPE7510) using high-pass cutoffs of 10-17 kbp depending on library length. Size-selected libraries were pooled for sequencing on the Revio platform using the Revio SPRQ Polymerase Kit (103-520-100) and Revio SPRQ sequencing plates (103-504-900) on Revio SMRT Cells (102-202-200) with Adaptive Loading and 30-hour movie times.

Saliva DNA were obtained using two extraction workflows: (1) Chemagic Magnetic Beads: DNA were previously extracted from saliva samples collected using DNA Genotek saliva collection devices. 600 ul of saliva was extracted using Chemagic MSM1 or Chemagic 360 DNA extraction instruments and eluted into TE buffer; and (2) Nanobind CBB Kit: A Nanobind CBB kit was used to collect HMW DNA from 500 uL saliva in DNA Genotek Oragene devices. DNA was extracted utilizing a Nanobind disk and stored in a low TE buffer. Extracted DNA were banked and later transferred to Broad Clinical Labs (BCL) for library preparation and sequencing. For circular consensus sequencing (CCS) library preparation, at least 2 μg of HMW genomic DNA were depleted of fragments <10 kbp using the PacBio SRE HT kit (103-124-500). The DNA were then sheared to ~15 kbp on the Hamilton Starlet system using automated pipette shearing. SMRTbell libraries were prepared using the PacBio HiFi Prep Kit 96 (103-122-600), including post-shear cleanup, DNA end repair, PacBio adapter ligation, and nuclease treatment. Libraries were subsequently hybridized with the PacBio standard sequencing primer and bound to SPRQ sequencing polymerase using the Revio SPRQ Polymerase Kit 96 (103-497-000). The resulting polymerase-bound SMRTbell complexes were normalized to 300 pM and pooled to achieve the target coverage (20X). CCS was performed on Revio instruments using 25M SMRT Cells (102-202-200) and Revio SPRQ Sequencing Plates (103-504-900), with a 2-hour pre-extension time and 30-hour movie time per SMRT cell.

HiFi reads were evaluated for nonhuman contamination using Kraken2^24^ (v2.1.3) and back-reference-qc pipeline (https://github.com/EichlerLab/back-reference-qc)^11^. The human-derived genomic reads were aligned to GRCh38 for male samples and to GRCh38noY (GRCh38 with the Y chromosome masked) for female samples using pbmm2 (v1.13.1, https://github.com/PacificBiosciences/pbmm2). Family pedigree, relatedness, and sample identity were confirmed, and potential cross-sample contamination was assessed, using VerifyBamID^70^ (v2.0.1), ntsm^71^ (v1.2.1), and Somalier^72^ (v0.2.19).

### Phased genome assembly construction

The HiFi assembly was constructed by hifiasm^25^ (v0.25.0). Parental WGS short or long reads were processed with yak (v0.1, https://github.com/lh3/yak.git) and then hifiasm trio-binning mode was used for phasing children samples, while the parental assemblies were partially phased by default. The sex chromosome contigs for father samples were corrected to hap1 for paternal haplotype (Y chromosome) and hap2 to maternal haplotype (X chromosome). N50 values were calculated and assembly QVs were evaluated by Merqury^73^ (v1.3) with k-mers from HiFi data (meryl^73^, v1.4) (**Dataset S1**).

### Rare variant discovery

SVs were called using the phased assembly variant, PAV^26^ caller (v2.4.6) and five alignment-based SV callers: PacBio SV calling and analysis tool (PBSV, v2.9.0, https://github.com/PacificBiosciences/pbsv), Sniffles^27^ (v2.3.2), DELLY^29^ (v1.3.3), Sawfish^30^ (v0.12.4), and cuteSV^28^ (v2.1.1). A validated SV callset was generated for each sample using Truvari^31^ (v5.2.0) with ‘truvari collapse’ and the following parameters: ‘--sizemin 50 --sizemax 100000 --gt het -k maxqual --intra --pctseq 0.8 --pctsize 0.8 --refdist 1000’. The validated callsets were then merged with SVs from 1,218 controls from Lin et al.^32^ using the following parameters: ‘--sizemin 50 --sizemax 100000 --keep maxqual --pctseq 0.9 --pctsize 0.9--refdist 500’ (**Dataset S2**). Rare SVs specific to probands were extracted from the collapsed callset using rareSVpool.py^11^ (v3) with ‘--type pro-only’ and subsequently filtered against gnomAD (v4.1.0) SVs using ‘truvari bench’ with the following parameters: ‘--pctsize 0.5 –pctseq 0.0 --pctovl 0.5 --sizefilt 50 --sizemax 100000’. The remaining SVs were annotated with comREG^11^ (v2.0.0) (**Dataset S3**), and the transmission status for pathogenic candidates was curated following the workflow described in Sui et al.^11^. Curation incorporated support from the alignment-based SV callers described above, genotyping with Kanpig^40^ (v2.0.2), tandem-repeat genotyping with TRGT^74^ (v1.4.1), and read- and quality-level evidence assessed using BoostSV^11,32^ (v1.0), subseq^26^, and the Integrated Genomics Viewer^75^ (IGV, v2.16.0).

Single-nucleotide variants and small indels were called for all 362 samples using DeepVariant^34^ (v1.10.0) from HiFi read alignments, followed by joint genotyping with GLnexus^35^ (v1.4.1). DNM candidates in 159 children were identified based on variants meeting the following criteria: a read depth (DP) of ≥10 across all family members (DP >4 for the male’s X chromosome), zero alternate reads in both parents (GT = 0/0 and AD = 0/0), and a genotype quality (GQ) of >20 and allele balance (AB) of >0.25 in the child. Variants in homopolymer regions, pseudoautosomal regions, centromeres, assembly gaps, and the highly repetitive Yq12 heterochromatic region were excluded. Proband DNMs (**Dataset S4**) that were absent from unaffected samples and the gnomAD v4.1.0 joint dataset were prioritized for annotation using VEP^37^ (v116.2) and comREG^11^ (v2.0.0).

Sequences encompassing potential pathogenic variants (**Dataset S5**) were extracted from assemblies using rustybam (v0.1.29, https://github.com/mrvollger/rustybam) and SAMtools^76^ (v1.19). Extracted sequences were aligned using minimap2^77^ (v2.26) and visualized across family haplotypes using the SVbyEye R package^78^ Multiple-sequence alignments were generated using MAFFT^79^ (v7.525) and visualized using Jalview^80^ (v2.11.4.1).

## Ethics

Ethical approval for this study was granted by the University of Washington IRB Committee B, under STUDY IDs: STUDY00000383 and STUDY00004522.

## Data availability

The underlying sequencing data, as well as the processed files used for analysis in this study are available to approved researchers through SFARI Base under Dataset IDs DS0000104 and DS0000134, and through the National Institute of Mental Health Data Archive (NDA) under Collection ID 3780.

## Code availability

Code and scripts used for the analyses presented in this manuscript are available in GitHub at https://github.com/EichlerLab/asap, under the MIT license. Any additional information required to reanalyze the data reported in this work paper is available upon request.

## Acknowledgments

We are grateful to all of the families at the participating Simons Simplex Collection (SSC) sites, as well as the principal investigators (A. Beaudet, R. Bernier, J. Constantino, E. Cook, E. Fombonne, D. Geschwind, R. Goin-Kochel, E. Hanson, D. Grice, A. Klin, D. Ledbetter, C. Lord, C. Martin, D. Martin, R. Maxim, J. Miles, O. Ousley, K. Pelphrey, B. Peterson, J. Piggot, C. Saulnier, M. State, W. Stone, J. Sutcliffe, C. Walsh, Z. Warren, E. Wijsman). We are also grateful to all of the families in Simons Foundation Powering Autism Research for Knowledge (SPARK), the SPARK clinical sites, and SPARK staff. We appreciate obtaining access to SSC phenotypic (DS0000071) and genetic (DS0000041) data, and SPARK phenotypic (DS0000035) and genetic (DS0000134, DS0000047) data through SFARI Base. Approved researchers can obtain the SPARK (DS0000035) and SSC (DS0000071) population datasets described in this study by applying at https://base.sfari.org. We gratefully acknowledge A. Wong, J.Hall, and A. Packer of the Simons Foundation, and P. Feliciano and J. Wright of the SPARK team at Boston Children’s Hospital, for their contributions and support. Thank you to the Simons Foundation for access to recruit participants through the SPARK Research Match program. We also thank Tonia Brown for assistance with manuscript editing. During the preparation of this work, the authors used ChatGPT to refine the language of the manuscript and Claude to assist with visualization code. Following the use of these tools, the authors reviewed and edited the content as needed and take full responsibility for the content of the publication. This work was supported, in part, by the US National Institutes of Health (NIH R01MH101221 to E.E.E.) and the Simons Foundation (SFI-AN-AR-Sex Differences-00017956-02 to E.E.E.). The content is solely the responsibility of the authors and does not necessarily represent the official views of the NIH. This work was also supported, in part, by the Brain Science and Brain-like Intelligence Technology-National Science and Technology Major Project (2025ZD0218000) and the National Natural Science Foundation of China (82471194) to T.W. E.E.E. is an investigator of the Howard Hughes Medical Institute.

## Author contributions

Y.S. and E.E.E. conceptualized the comparative analysis of blood-derived LRS data from the University of Washington and saliva-derived LRS data from SPARK. K.H. and E.N. coordinated sample collection. K.M.M., K.H., and M.A. generated the sequencing data. Y.S., Y.K., I.W., and N.K. performed data quality control. Y.S. conducted the formal analyses, developed the methodology, and created the visualizations. G.C. and T.W. performed validation and phenotypic investigation. Y.S. and E.E.E. wrote the original draft. Y.S., E.E.E., G.C., T.W., Y.K., I.W., N.K., M.A., K.H., K.M.M., and E.N. reviewed and edited the manuscript. E.E.E. supervised the study and provided funding.

## SUPPLEMENTAL INFORMATION

### Supplemental Datasets S1-S5 (see separate files)

Dataset S1. Sample manifest and data production

Dataset S2. Collapsed SVs in 362 and 1,218 controls

Dataset S3. Rare SVs in 107 probands

Dataset S4. Private DNMs in 102 probands

Dataset S5. Summary of pathogenic and candidate variants

